# Cognitive Impairment and Noncommunicable Diseases in Egypt’s Aging Population: Insights and Implications from the pilot of the Longitudinal Study of Egyptian Healthy Aging “AL-SEHA”

**DOI:** 10.1101/2023.10.16.23297117

**Authors:** Sara A. Moustafa, Reem Deif, Nada Gaballah, Mohamed Salama

**Affiliations:** Institute of Global Health and Human Ecology, The American University in Cairo; Department of Computer Sciences, School of Sciences and Engineering, The American University in Cairo; Global Brain Health Institute (GBHI), Trinity College Dublin (TCD), Dublin, Ireland

## Abstract

As the global population ages, the prevalence of cognitive impairment among older individuals has been steadily rising. Egypt, like many countries, is grappling with the challenges posed by an aging demographic. Cognitive impairment not only affects the quality of life of older adults but also imposes significant burdens on healthcare systems and societies as a whole. This paper presents findings from the pilot phase of the Longitudinal Study of Egyptian Healthy Aging, known as “AL-SEHA,” shedding light on the intricate relationship between cognitive impairment and noncommunicable diseases (NCDs) in Egypt’s aging population.

**KEY MESSAGES:**

- The increasing prevalence of cognitive impairment among the aging population creates a need for a comprehensive understanding of its implications on the individuals’ quality of life
- A significant association between cognitive impairment and non-communicable diseases exists and further research can help in the strategic allocation of resources for designing the most effective interventions

## Introduction

The concept of healthy aging, as advocated by the World Health Organization, emphasizes nurturing functional ability in older age for an enriched quality of life. This approach integrates intrinsic capacity (IC) and environmental factors, transitioning elder care from disease-centered to proactive personalized interventions, notably in hospitalized older adults^1^. The innovative notion of healthy aging encapsulates the process of nurturing and sustaining functional ability in older age for an enriched quality of life. This perspective underscores the essence of functional ability, encompassing health-related attributes that empower older individuals to pursue valued endeavors. It intertwines an individual’s (IC), a composite of physical and mental capabilities, with environmental factors, thereby enhancing the potential for a higher quality of life while reducing societal burdens. Traditional elder care approaches, centered on specific disease markers, are transitioning towards a longitudinal, proactive model driven by personalized interventions to amplify intrinsic capacity and functional ability, particularly notable in the context of hospitalized older adults ^1,2^.

### Assessing Intrinsic Capacity and Predicting Health Outcomes

The assessment of IC’s indicators predicts health outcomes, particularly functional capability. Domains such as locomotion, vitality, cognition, psychological, and sensory elements are pivotal for maintaining IC in older adults ^3^. Beard’s study ^4^ corroborates this, identifying five subfactors within the English Longitudinal Study on Ageing (ELSA) that forecast future functioning. However, a universally accepted IC index is yet to emerge, warranting further exploration. The significance of assessing IC becomes evident in its predictive value for health outcomes, specifically concerning functional capability. The intricate nature of IC indicators raises inquiries about the most pertinent markers for comprehensive evaluation of an individual’s holistic physical and mental condition. Recent Chinese Longitudinal study CHARLS further substantiates this structural validity through the ELSA approach ^5^, collectively accentuating the predictive potential of IC decline and its domains within aging populations. However, a universally acknowledged IC index for clinical or research purposes is yet to be established, necessitating further exploration and quantification of IC concepts in varied contexts.

### Cognitive Impairment’s Impact on Quality of Life

Cognitive impairment’s escalating prevalence, especially with age, necessitates a comprehensive exploration of its implications for individuals’ quality of life ^6,7^. The escalation in cognitive decline, particularly with increasing age and life expectancy, mirrors the mounting challenges facing developed and developing nations. The resultant surge in dementia not only inflicts substantial economic and social burdens on patients and families but also warrants a comprehensive exploration of its prevalence across genders and residential areas. Discrepancies within the existing literature concerning the gender and residential area-based prevalence of cognitive impairment highlight the need for a cohesive understanding of these dimensions ^6,7^.

### Socioeconomic Factors and Cognitive Impairment Risk

Socioeconomic factors intricately interplay with cognitive health, with lower education, job insecurity, and lifestyle choices influencing susceptibility to cognitive impairments ^9^. Notably, individuals with lower educational attainment, diminished IQ, and tenuous job security exhibit higher susceptibility to cognitive impairments. Furthermore, lifestyle choices play a pivotal role, with tobacco and alcohol usage surfacing as significant predictors of cognitive decline. These connections emphasize the multifaceted nature of cognitive health, woven intricately with socioeconomic dimensions and behavioral patterns.

### Noncommunicable Diseases (NCDs) and Cognitive Decline

NCDs, like diabetes and hypertension, impact cognitive faculties, revealing the interconnectedness of physical and cognitive well-being (World Health Organization, 2016)^10^. NCDs, a formidable presence within the global health landscape, assume a central role in shaping morbidity and mortality patterns. Their impact resonates through reduced problem-solving capabilities, compromised verbal proficiency, and waning numerical comprehension, underscoring the intricate interplay between physical and cognitive well-being^10^.

### Cognitive Impairment and NCDs in Egypt: Gaps in Literature

As people age, they are more likely to experience chronic non-communicable conditions. Based on the recommendations of WHO, the National Institute of Aging (NIA), and the Centers for Disease Control and Prevention (CDC), fighting non-communicable diseases is broadly agreed to be the key to father health gains at an older age and to making health and social policies sustainable. NCDs, malnutrition, and depression are common in our elderly population, particularly among women and the under-educated ^11^.

Aging is often accompanied by cognitive function, memory, and mental health declines. Research on effective interventions to prevent or delay these declines and improve the quality of life of older adults is critical ^12^. With a prevalence of depression, a sense of isolation, and declined cognitive functions in the Egyptian elderly ^12,13^. In older people, cognitive impairment is a strong predictor of functional disability and the need for care. Mild cognitive impairment increases the risk of developing dementia, and available evidence suggests that a five-year delay in the age of onset would reduce dementia prevalence by half ^14,15,16^. There is limited data on mental health and cognitive functions in the Arab world, and specifically in Egypt where data are specific to certain areas or specific clinical populations rather than those derived from large nationwide studies. With this age group expected to grow by 2050, the consequences of dementia will grow as well.

Existing research often focuses on cognitive impairment within NCDs, leaving a gap in understanding cognitive health among the elderly holistically. Extant investigations into cognitive impairment have predominantly focused on individuals afflicted by NCDs, imparting a limited scope to understanding the broader landscape of cognitive health among the elderly. Prior research has, in various instances, examined the intersection of cognitive issues and NCDs, potentially excluding a more holistic comprehension of cognitive health among the elderly population. Addressing this void, our study endeavors to illuminate the prevalence of cognitive impairment within Egypt’s older demographic, specifically those grappling with chronic diseases.

### Investigating Cognitive Impairment Among Egypt’s Older Population

## Methods

Preliminary results of the pilot phase of the Egyptian Longitudinal Study for Aging (AL-SEHA), an HRS sister study adopting the SHARE questionnaire presented in Table 3, offer insights into Egypt’s aging demographic. The pilot study includes respondents aged 50 and above, with a sample size of 299 participants. The results reflect a diversity of conditions, with varying percentages of cognitive impairment across diseases. Responding to this need, an Egyptian aging study modeled after the Health and Retirement Survey (HRS) network studies is being launched. Our findings are based on the AL-SEHA questionnaire, which adapted the SHARE survey, targeting noninstitutionalized individuals aged 50 and above ^17^. This encompassing longitudinal survey included all those above 50 within chosen households.

The studies involving human participants were reviewed and approved by IRB of the American University in Cairo (IRB-AUC). The patients/participants provided their written informed consent to participate in this study. The current work has been approved by the IRB of the American University in Cairo. IRBAUC_ Case# 2021-2022-029

### Research design

The study used a cross-sectional design, assessing both dependent and independent variables simultaneously. Findings were based on a questionnaire from the Longitudinal Study of Egyptian Healthy Aging (AL-SEHA), adapted from the European Survey of Health, Ageing, and Retirement (SHARE).

Interviews included individuals aged 50 and above, as well as eligible partners living in the same household. Some questionnaire modules were not given to all household respondents, and interview order within couples was random.

### Inclusion criteria

The study included individuals who could communicate and lived independently at home, in rented accommodation, hostels, or retirement homes. Those with severe psychiatric issues were excluded. The sample size was 299 participants, selected through a non-probability convenience sampling technique based on the eligibility criteria.

### Fieldwork

Investigators from five universities were trained to collect data using an Arabic-translated SHARE form. A sample of eligible elderly individuals was chosen, and home visits were conducted to interview 15 participants per investigator. Data collection took about a month and was uploaded to the American University in Cairo’s Social Research Centre’s internet server.

### Demographic factors

AL-SEHA data set contains detailed data on demographics. Summary statistics of all demographic variables used in the analyses can be found in Table 1.

**Table 1.** Demographics.

|  | N | % |
| --- | --- | --- |
| **Gender** |  |  |
| Male | 128 | 42.81 |
| Female | 171 | 57.19 |
| **Educational Attainment** |  |  |
| No education | 48 | 16.05 |
| Primary education | 31 | 10.37 |
| Preparatory education | 18 | 6.02 |
| High school | 48 | 16.05 |
| Medium high | 18 | 6.02 |
| Bachelor | 112 | 37.46 |
| Post grad | 24 | 8.03 |
| **Age** |  |  |
| 50-60 | 151 | 50.5 |
| 60-70 | 88 | 29.43 |
| 70-80 | 50 | 16.72 |
| 80-90 | 8 | 2.68 |
| Above 90 | 2 | 0.67 |
| **Marital Status** |  |  |
| Unmarried | 5 | 1.67 |
| Engaged | 0 | 0 |
| Married | 190 | 63.55 |
| Separated | 3 | 1 |
| Divorced | 13 | 4.35 |
| Widow | 88 | 29.43 |

### Cognitive function

The data were extracted from the SPSS (Version 25) dataset for statistical analysis. Bivariate analyses included chi-square tests. As appropriate, data were compared using independent t-tests, presented in Table 2.

**Table 2.** Cognitive functions.

|  | Impaired (n=118) |  | Unimpaired (n=181) |  | Chi Squared Test | p-value |
| --- | --- | --- | --- | --- | --- | --- |
| **Age** | N | % | N | % |  |  |
| 50-60 | 52 | 44.07 | 99 | 54.7 |  |  |
| 60-70 | 39 | 33.05 | 49 | 27.07 |  |  |
| 70+ | 27 | 22.88 | 33 | 18.23 | 3.23 | 0.2 |
| Mean (SD) | 62.338(8.8087) |  | 60.635(8.0505) |  |  |  |
| **Gender** |  |  |  |  |  |  |
| Male | 44 | 37.29 | 84 | 46.41 |  |  |
| Female | 74 | 62.71 | 97 | 53.59 | 2.07 | 0.15 |
| **Education** |  |  |  |  |  |  |
| No education | 29 | 24.58 | 19 | 10.5 |  |  |
| Basic/intermediate | 52 | 44.07 | 63 | 34.81 |  |  |
| University | 37 | 31.36 | 99 | 54.7 | 18.968 | **0.0001** |
| **Marital Status** |  |  |  |  |  |  |
| Single | 1 | 0.85 | 4 | 2.21 |  |  |
| Married | 69 | 58.47 | 121 | 66.85 |  |  |
| Separated | 3 | 2.54 | 0 | 0 |  |  |
| Divorced | 3 | 2.54 | 10 | 5.52 |  |  |
| Widow | 42 | 35.59 | 46 | 25.41 | 10.159 | **0.0378** |

### Dependent variable

Cognitive performance was assessed using a composite measure based on three tests: immediate recall, delayed recall, and verbal fluency. The immediate and delayed recall tasks evaluated short-term verbal learning and memory. Verbal fluency measured executive function and language ability. The composite score ranged from 0 to 1, where 0 to 0.5 indicated impairment and 0.5 to 1 indicated unimpaired cognitive function.

### Covariates and moderators

Cognitive performance in later life is influenced by various factors such as age, education, gender, and marital status. To account for these variables, they were included as covariates in the analyses. Gender was represented as a dummy variable with 1 for men and 2 for women. Age was also represented as a dummy variable with 1 for ages between 50 and 60, 2 for ages between 60 and 70, and 3 for ages over 70. Similarly, education was represented as a dummy variable with 1 for no education, 2 for basic to intermediate education, and 3 for university or postgraduate studies.

### Analyses of data

To begin, descriptive statistics were used to compute The means and standard deviations of continuous variables, as well as the percentages and frequencies of categorical variables.

Bivariate analyses were performed in the second stage to examine the relationship between cognitive health and the study variables, using independent t-tests for factors.

### Cognitive performance

The researchers assessed cognitive functioning using three tasks: verbal fluency, immediate word recall, and delayed word recall. Verbal fluency evaluated executive function by asking participants to name as many animals as possible in one minute. Memory function was assessed through the Ten-Word Delayed Recall Test, where participants recalled ten words immediately and after five minutes. These measures allowed researchers to evaluate executive function and memory constructs, differentiating between age-related memory changes and significant impairments. The assessment was based on widely-used neuropsychological tasks for memory performance evaluation.

### Prevalence of Noncommunicable Diseases (NCDs)

Our study focuses on cardiovascular, abdominal, chest, neuro/psychiatric, metabolic, and musculoskeletal diseases and cancer’s impact on cognitive impairment presented in Table 3.. The prevalence of these diseases varies across age groups and genders, reflecting the intricate web of interactions shaping Egypt’s aging population.

**Table 3.**
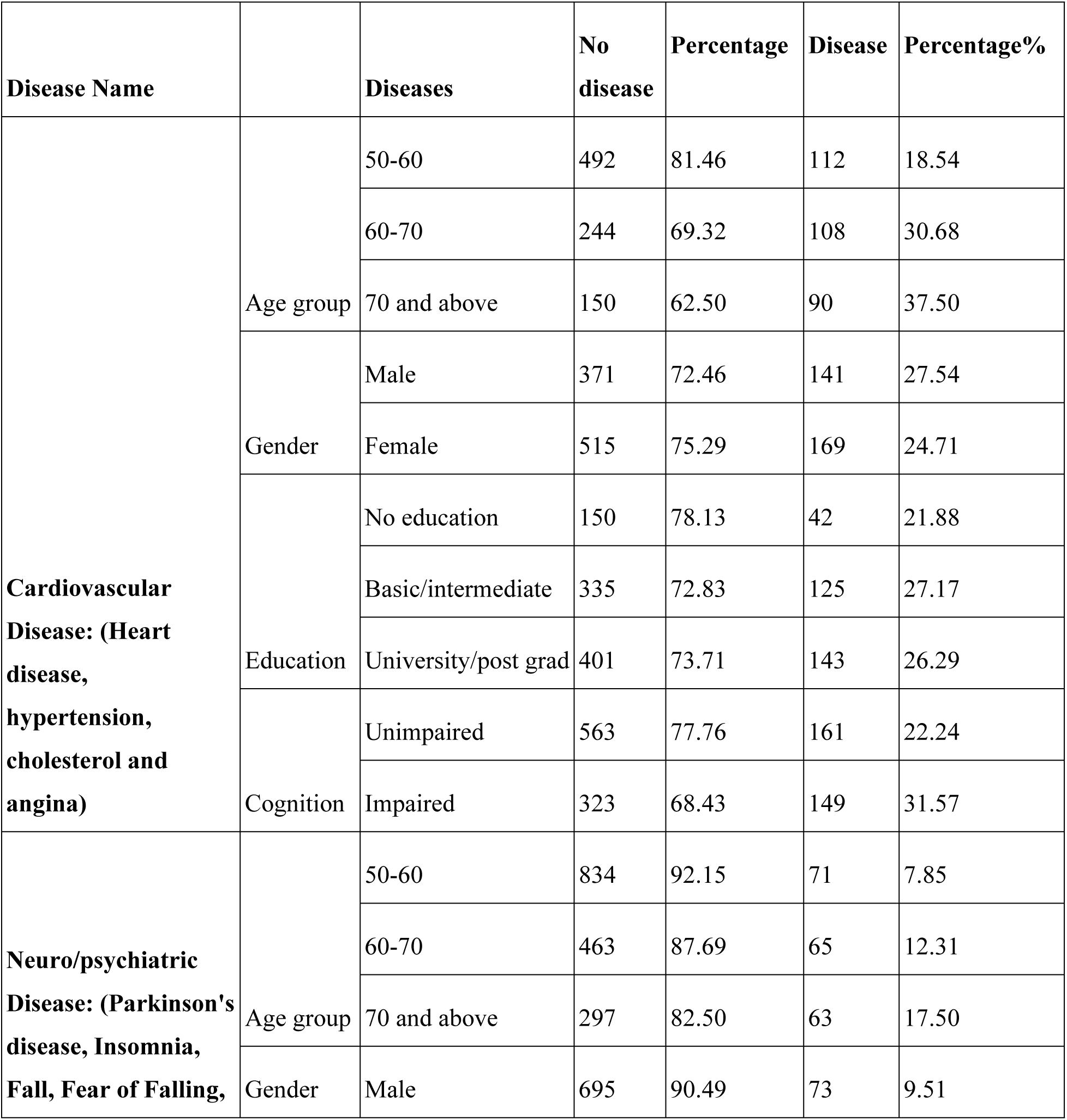
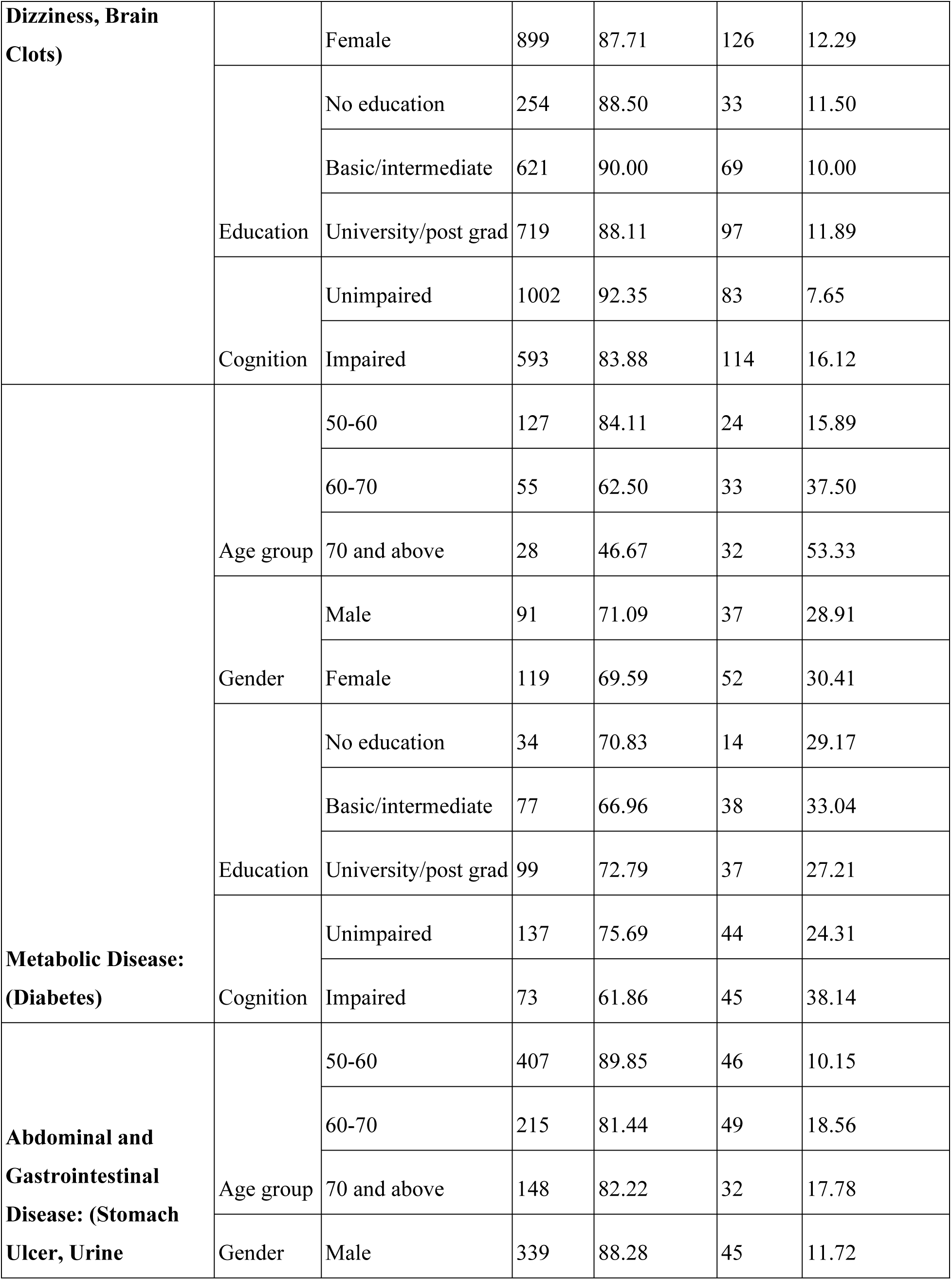
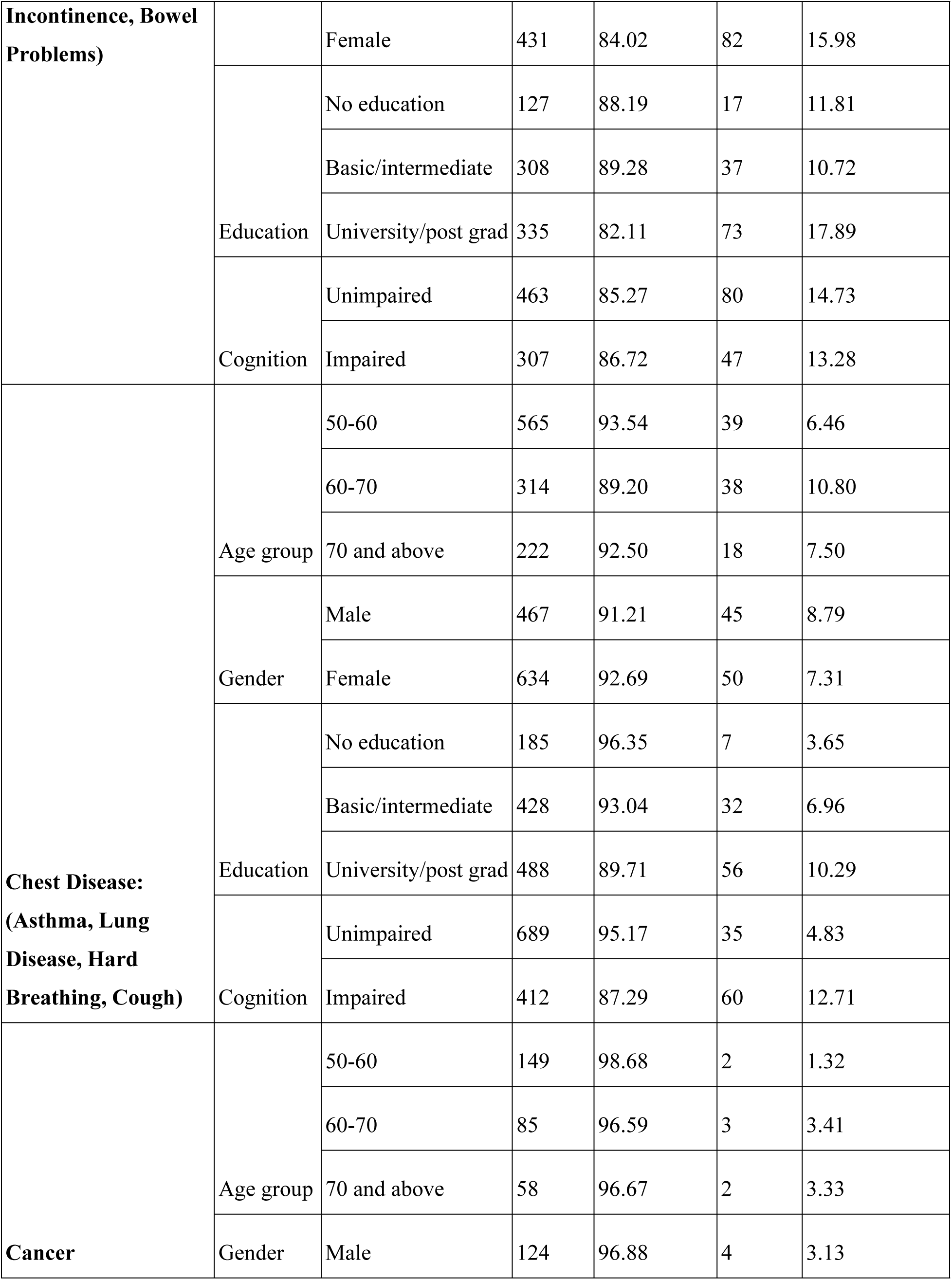
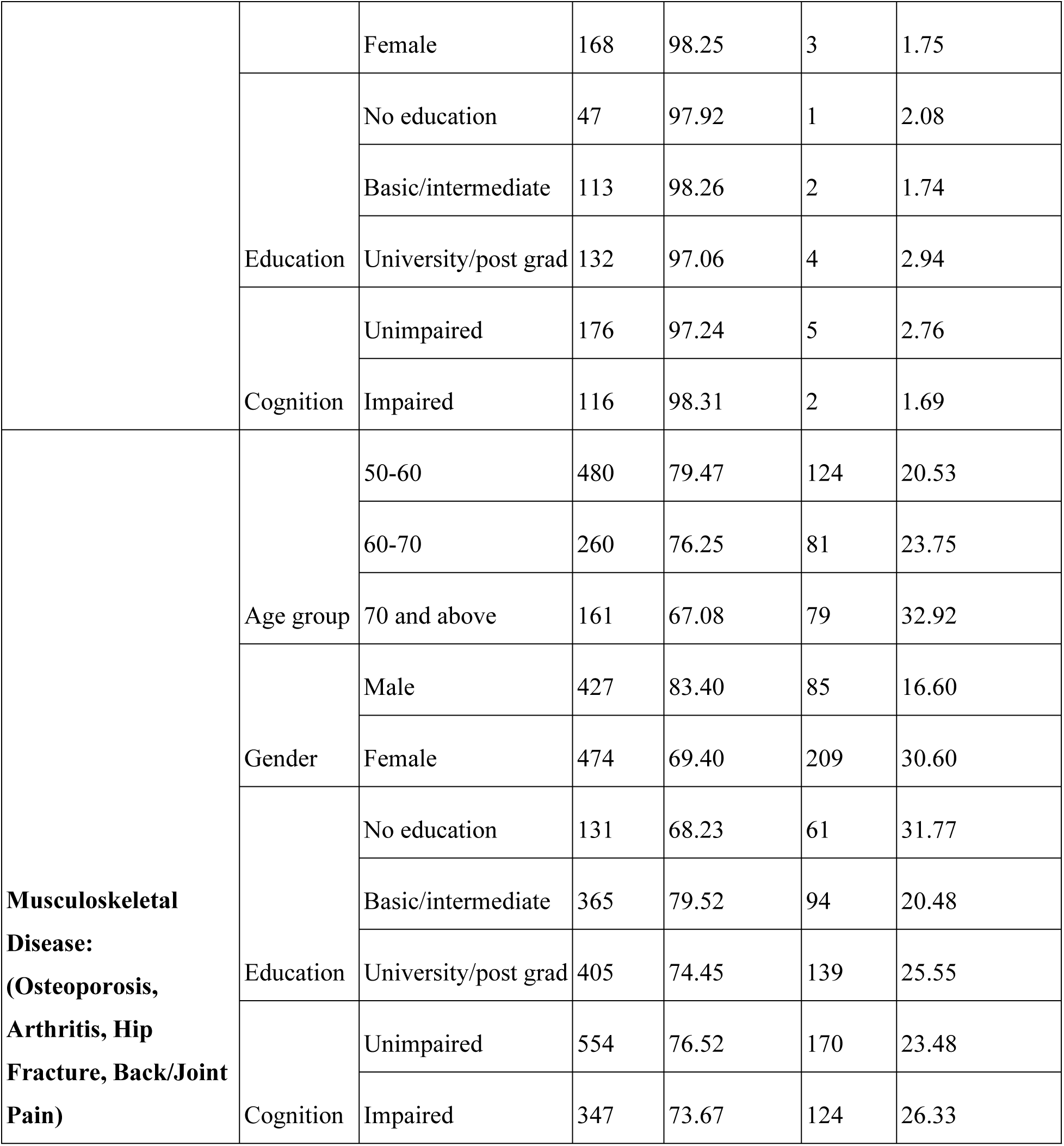
Non-communicable diseases and cognitive functions preliminary results.

### Neuro/Psychiatric Diseases and Cognitive Well-being

Cognitive impairment reveals the interconnectedness of mental well-being and cognitive faculties. Conditions like insomnia show statistically significant associations with cognitive impairment (p-value: 0.0169). The intricate interplay between cognitive well-being and mental health becomes evident when exploring associations between neuro/psychiatric diseases and cognitive impairment.

### Metabolic Diseases and Their Impact on Cognition

Metabolic diseases, epitomized by diabetes, demonstrate their nuanced relationship with cognitive health through statistically significant associations with cognitive impairment (p-value: 0.0137). These findings accentuate the urgency of interventions that holistically address metabolic and cognitive well-being, thereby potentially ameliorating the progression of cognitive decline.

### Respiratory Health and Cognitive Implications

Associations between chest diseases and cognitive impairment underline the intricate connection between respiratory health and cognitive well-being. Conditions like hard breathing and cough show statistically significant associations with cognitive impairment (p-values: 0.000076, 0.0032). The intricate correlation between respiratory health and cognitive well-being materializes in associations between chest diseases and cognitive impairment.

### Musculoskeletal Health and Functional Independence

The intricate dance between physical well-being and cognitive health manifests as associations between musculoskeletal diseases and cognitive impairment. Ailments like falls, fear of falls, and dizziness, often rooted in musculoskeletal health, demonstrate statistically significant associations with cognitive impairment, underscoring the profound interaction between the physical and cognitive domains (p-values: 0.00108, 0.001089, 0.0089).

### Cancer and Its Nuanced Prevalence

Cancer prevalence in the older Egyptian population was notably low. Specifically, among those aged 50- 60 years, only 1.32% had cancer, with 98.68% without the disease. In the 60-70 age group, 3.41% were affected by cancer, while 96.59% were not. Similarly, among individuals aged 70 and above, cancer was found in 3.33%, with 96.67% unaffected. However, Larger and more diverse samples are needed to draw more definitive conclusions about cancer rates in the Egyptian older population. Acknowledging these limitations is essential for a balanced perspective on the findings, and further research is required for more accurate insights into the cancer burden among the older population in Egypt.

## Discussion

### Implications for Egypt’s Aging Population

Findings hold significance for Egypt’s aging demographic, informing policies and interventions that cater to older adults’ unique needs. The implications of these findings extend far beyond statistical associations. They reverberate through policy decisions, healthcare interventions, and societal strategies that acknowledge the diverse needs and challenges faced by Egypt’s aging demographic. These implications underscore the imperatives of tailored interventions that holistically address the intricate interplay between physical and cognitive well-being.

The policy implications derived from the AL-SEHA pilot study data present a comprehensive framework for the development of effective strategies aimed at addressing noncommunicable diseases (NCDs) and cognitive health concerns within Egypt’s elderly population. To address these issues, it is imperative to craft and implement targeted programs with a strong focus on the prevention and management of NCDs, with particular attention to the promotion of cognitive health and overall well-being. Additionally, the evaluation of the necessity for disease prevention initiatives, including adult vaccination campaigns, awareness programs addressing tobacco use, and initiatives that encourage physical activity among older adults, is crucial. Furthermore, enhancing the geriatric care landscape in Egypt is essential, involving the incorporation of specialized care pathways that cater to the unique needs of the elderly. This also necessitates a revision of the medical education curriculum to include geriatric medicine, emphasizing the diagnosis, treatment, and management of multiple coexisting health conditions prevalent in the aging population. Expanding the capacity and accessibility of geriatric care facilities is vital to ensure comprehensive and effective support for older individuals. Furthermore, the integration of geriatric healthcare services across primary, secondary, and tertiary healthcare systems, including community-based health initiatives, is essential to provide holistic care. Cross-regional analyses should be conducted to identify best practices in areas such as long-term care, home-based support, legal protections, and healthcare expenditure, customized to the specific context of Egypt. Additionally, the formulation of national strategies for dementia prevention, targeting both the general population and vulnerable subgroups, is paramount. This should be complemented by the establishment of governmental initiatives at both national and local levels to effectively address the multifaceted challenges of dementia care and provide support for patients and caregivers in Egypt. These policy recommendations collectively form a robust foundation for addressing the pressing healthcare needs of Egypt’s elderly population in the context of NCDs and cognitive health.

### Healthcare Planning and Resource Allocation

Understanding associations between cognitive impairment and diseases aids in strategic resource allocation for timely interventions. The recognition of associations between cognitive impairment and diverse diseases augments the strategic blueprint for resource allocation within the Egyptian healthcare system. Informed by these associations, policymakers can channel resources towards timely interventions that ameliorate cognitive decline, enhancing not only individual well-being but also alleviating societal burdens.

### Catalyzing Research and Collaboration

Associations between cognitive impairment and diseases foster interdisciplinary research and collaboration for holistic strategies. The implications of cognitive impairment’s associations with diseases extend beyond individual interventions, catalyzing collaborative research across disciplines in Egypt and the MENA region. This interdisciplinary endeavor propels the creation of comprehensive strategies that transcend traditional healthcare silos, forging innovative pathways to address the intricate web of cognitive decline.

### Conclusion: Enabling a Brighter Aging Future for Egypt

Evidence-based policies and interventions pave the way for a healthier and more inclusive aging demographic. In conclusion, the synthesis of evidence-based policies and interventions emerges as the cornerstone of facilitating a brighter, healthier aging trajectory for Egypt. The intricate interplay between cognitive impairment and noncommunicable diseases unravels novel avenues for policymaking and interventions, steering the nation toward a future that prioritizes the well-being and dignity of its aging population.

### Limitations of the Study and Suggestions for Future Research in the Egyptian Context

Acknowledging limitations, future research can employ longitudinal designs and objective measures to enhance understanding. Admitting the inherent limitations of this study, future research endeavors within the Egyptian context can harness the power of longitudinal designs and objective measurements. These methodological refinements promise to illuminate deeper insights into the dynamic interplay between cognitive impairment and diseases, thereby enriching the foundation for comprehensive interventions.

In conclusion, this paper delved into the intricate web connecting cognitive impairment and noncommunicable diseases within Egypt’s aging population. By investigating associations between cognitive decline and diverse diseases, we illuminated a tapestry of interactions that transcend traditional healthcare boundaries. The implications of these findings resonate through evidence-based policies, cultural considerations, interdisciplinary collaboration, and most importantly, empowered aging experiences. These insights paves the way for Egypt towards a future characterized by enhanced well-being, dignity, and inclusivity for its elderly citizens.

## Data Availability

All data are fully available and without restrictions

## Acknowledgment

The present work has been supported by the German Academic Exchange Services (DAAD), through the funding program: Higher Education Dialogue with the Muslim World, project (Ageing in the East Mediterranean Region: EMage). This work benefitted from an incentive grant from the World Health Organization (WHO) as part of the WHO Global Noncommunicable Disease (NCD) Platform Young Researchers Programme (Grant reg. № 2022/1256080).

The authors want to thank Dr. Tea Collins and Ms. Daria Berlina for their support and feedback on the final draft.

## Patient involvement

No patients were involved.

